# Determinants of Caregiver Well-Being in End-of-Life Care: A Systematic Review Protocol

**DOI:** 10.1101/2025.06.23.25330162

**Authors:** Yu Xuan NG, Rui Fang Teo, Ravi Shankar

## Abstract

End-of-life caregiving represents one of the most demanding experiences for informal caregivers who often sacrifice their own well-being while supporting dying loved ones. This systematic review protocol presents a comprehensive framework for synthesizing evidence on factors influencing caregiver well-being in end-of-life care settings. Using the SPIDER framework (Sample, Phenomenon of Interest, Design, Evaluation, Research type), we will investigate caregiver experiences across physical, psychological, social, and spiritual well-being dimensions. Eight electronic databases (PubMed, Web of Science, Embase, CINAHL, MEDLINE, Cochrane Library, PsycINFO, Scopus) will be searched from inception to June 2025. Eligible studies include quantitative, qualitative, and mixed-methods research examining factors associated with caregiver well-being in hospice, palliative, and terminal care contexts. Covidence software will facilitate systematic screening, data extraction, and quality assessment by two independent reviewers using design-specific appraisal tools. Data synthesis will employ a convergent integrated approach combining narrative synthesis with meta-analysis where appropriate. Quality assessment will utilize JBI checklists, Newcastle-Ottawa Scale, and CASP tools, with evidence certainty evaluated using GRADE and GRADE-CERQual. This protocol follows PRISMA-P guidelines and will be registered with PROSPERO. Findings will inform evidence-based interventions and policies supporting the millions providing essential end-of-life care.

## Introduction

The landscape of end-of-life care has transformed dramatically, with over 40 million people requiring palliative care annually worldwide (World Health Organization, 2020). Informal caregivers—predominantly family members but also friends and volunteers— provide 70-80% of care received by terminally ill individuals, contributing economic value worth hundreds of billions annually to healthcare systems (National Alliance for Caregiving, 2020). The shift toward home-based end-of-life care, driven by patient preferences and healthcare policies, has intensified demands on informal caregivers who now perform complex medical tasks once reserved for professionals, including pain management, symptom monitoring, and medication administration (Bee et al., 2009).

Caregiver well-being encompasses multiple interconnected dimensions. Physical well-being includes maintaining energy, sleep quality, and functioning despite caregiving demands. Research documents alarming health deterioration among end-of-life caregivers, including increased cardiovascular disease and compromised immunity (Schulz & Beach, 1999). Psychological well-being extends beyond absence of symptoms to include life satisfaction, meaning, and personal growth. Meta-analyses indicate 30-40% of caregivers experience clinical depression and anxiety, substantially exceeding general population rates (Pinquart & Sörensen, 2003). Social well-being involves maintaining relationships and activities outside caregiving, often compromised by caregiving’s all-consuming nature (Harding et al., 2012; Methi et al., 2024). Spiritual well-being encompasses finding meaning and maintaining hope while facing loss (Applebaum et al., 2014; Karacan et al., 2025).

### Theoretical Framework

Multiple theoretical perspectives inform understanding of caregiver well-being determinants. The Stress Process Model conceptualizes caregiving through primary stressors (direct care demands), secondary stressors (spillover effects), mediating factors (coping, support), and outcomes (Pearlin et al., 1990). The Transactional Model emphasizes cognitive appraisal in determining stress responses and coping effectiveness (Lazarus & Folkman, 1984). Conservation of Resources Theory suggests well-being suffers when valued resources are threatened or lost, particularly relevant in end-of-life care’s rapid resource depletion (Hobfoll, 1989). Social Ecological Models recognize influences operating across individual, interpersonal, organizational, community, and policy levels (Bronfenbrenner, 1979).

### Evidence Gaps and Rationale

Despite substantial research on caregiver experiences, evidence remains fragmented across disciplines and populations. Previous reviews typically examined specific outcomes like burden or depression rather than comprehensive well-being determinants (Grande et al., 2009). Methodological limitations include inconsistent well-being conceptualization, convenience-based determinant selection, and underrepresentation of diverse populations including male caregivers, young adults, and ethnic minorities (Washington et al., 2012). The predominance of cross-sectional designs limits understanding of temporal relationships, while emphasis on negative outcomes neglects factors promoting positive adaptation (Collins & Swartz, 2011).

This systematic review addresses critical needs from multiple perspectives. Humanitarian imperatives recognize caregivers’ invaluable contributions deserve support. Healthcare quality depends on caregiver well-being, with stressed caregivers struggling to provide optimal care (Waldrop et al., 2005). Economic implications include lost productivity, increased healthcare utilization, and long-term health consequences (Van Houtven et al., 2013). Policy development requires evidence to inform resource allocation and support strategies.

### Objectives

This systematic review aims to comprehensively identify, evaluate, and synthesize evidence on caregiver well-being determinants in end-of-life care settings. Primary objectives include identifying factors associated with well-being across personal, relational, care-related, and systemic domains. Secondary objectives examine variation across populations and contexts, explore determinant interrelationships, evaluate evidence quality, and identify research gaps. Specific research questions address: (1) personal factors including demographics, health, personality, and coping; (2) relational factors including relationship quality, family dynamics, and social support; (3) care-related factors including duration, intensity, and patient needs; (4) environmental/systemic factors including healthcare systems, services, and cultural contexts; (5) pathways and mechanisms of influence; and (6) variations across subgroups and care trajectories.

## Methods

### Protocol Development and Registration

This protocol follows PRISMA-P 2015 guidelines ensuring comprehensive reporting (Moher et al., 2015). Development involved iterative refinement through team discussions, expert consultation, and pilot testing. The protocol is registered with PROSPERO (CRD42024599362) before commencing the review. Multiple frameworks guide the review: SPIDER for structuring questions about complex phenomena, PICOS for quantitative associations, and FINER criteria ensuring feasibility and relevance (Cooke et al., 2012; Hulley et al., 2013). Cochrane Handbook principles apply where appropriate, adapted for determinant-focused synthesis (Higgins et al., 2019).

### Eligibility Criteria

Population includes informal/unpaid caregivers providing physical, emotional, practical, or spiritual support to individuals receiving end-of-life care. Professional caregivers are excluded unless informal caregiver data can be extracted separately. End-of-life contexts encompass hospice, palliative, terminal care, and care for advanced life-limiting illnesses. The phenomenon of interest is caregiver well-being across physical, psychological, social, spiritual, and quality of life dimensions, with examination of associated determinants at any ecological level. Eligible designs include quantitative (cross-sectional, cohort, case-control), qualitative (all approaches), and mixed-methods studies. Intervention studies are included only for baseline associations. Studies must be peer-reviewed, published from inception to June 2025, and available in English. Exclusions include bereavement-only studies, patient-only outcomes, theoretical papers without empirical data, and grey literature.

**Table 1:** Inclusion and Exclusion Criteria.

| Criterion | Inclusion | Exclusion |
| --- | --- | --- |
| **Population** | • Informal/unpaid caregivers<br>• Any relationship to patient<br>• Any age or gender | • Professional caregivers only<br>• Bereaved caregivers only<br>• Non-end-of-life contexts |
| **Phenomenon** | • Caregiver well-being (any dimension)<br>• Determinants at any level | • No well-being measures<br>• Theoretical only |
| **Design** | • Quantitative, qualitative, mixed-methods<br>• Baseline intervention data | • Reviews, opinions, case reports<br>• Conference abstracts only |
| **Evaluation** | • Validated well-being measures<br>• Statistical/thematic analysis | • No extractable data |
| **Research Type** | • Empirical, peer-reviewed<br>• English language | • Grey literature<br>• Non-English (if not translatable) |

### Search Strategy

Eight databases ensure comprehensive coverage: PubMed (MEDLINE plus recent citations), Web of Science (multidisciplinary), Embase (European focus), CINAHL (nursing/allied health), MEDLINE via Ovid (sophisticated interface), Cochrane Library (systematic reviews/trials), PsycINFO (psychological literature), and Scopus (citation tracking). Strategy development involved health sciences librarian collaboration, pilot testing, and PRESS checklist peer review (McGowan et al., 2016). The search combines controlled vocabulary with free-text terms:

((“caregiver*” OR “carer*” OR “care giver*” OR “family caregiver*” OR “informal caregiver*” OR “family member*” OR “spouse*” OR “partner*” OR “relative*” OR “unpaid caregiver*”) AND (“end-of-life” OR “end of life” OR “terminal care” OR “hospice*” OR “palliative” OR “dying” OR “life-limiting” OR “advanced illness”) AND (“well-being” OR “wellbeing” OR “quality of life” OR “mental health” OR “physical health” OR “adjustment” OR “coping” OR “burden” OR “stress” OR “satisfaction”) AND (“determinant*” OR “factor*” OR “predictor*” OR “correlate*” OR “association*” OR “influence*”))

### Study Selection

Covidence software manages the selection process with complete audit trails (Veritas Health Innovation, 2021). Following training and calibration exercises achieving Cohen’s kappa ≥0.60, two reviewers independently screen titles/abstracts then full texts. Disagreements resolve through discussion or third reviewer arbitration. Exclusion reasons follow a hierarchy: wrong population, phenomenon, design, outcomes, or other. PRISMA flow diagrams document the process.

### Data Extraction

A standardized form captures: study characteristics (design, setting, sample), participants (demographics, relationships, caregiving duration/intensity), determinants (constructs, measures, timing), outcomes (well-being domains, instruments), and findings (associations, themes). Two reviewers extract independently with reconciliation of discrepancies. Authors are contacted for missing data with four-week response windows.

### Quality Assessment

Design-specific tools evaluate methodological quality. Cross-sectional studies: JBI Checklist examining inclusion criteria, measurement validity, confounding, and analysis (Moola et al., 2020). Cohort studies: Newcastle-Ottawa Scale assessing selection, comparability, and outcome ascertainment (Wells et al., 2021). Qualitative studies: CASP Checklist evaluating aims, methodology, design, recruitment, data collection, reflexivity, ethics, analysis, and findings (CASP, 2018). Mixed-methods: MMAT addressing qualitative, quantitative, and integration quality (Hong et al., 2018). Two reviewers assess independently with consensus resolution.

### Data Synthesis

Convergent integrated design enables meaningful integration across methodologies. Narrative synthesis organizes findings by determinant categories within the social-ecological model. Meta-analysis using random-effects models pools effect estimates where homogeneous, with I^2^ statistics assessing heterogeneity. Subgroup analyses explore variations by study/participant characteristics. Qualitative thematic synthesis involves line-by-line coding, descriptive theme development, and analytical theme generation (Thomas & Harden, 2008). Integration uses joint displays mapping quantitative and qualitative findings to identify convergence, divergence, and complementarity.

### Evidence Certainty

GRADE assesses quantitative evidence considering risk of bias, inconsistency, indirectness, imprecision, and publication bias (Guyatt et al., 2008). GRADE-CERQual evaluates qualitative findings through methodological limitations, coherence, adequacy, and relevance (Lewin et al., 2018). Summary tables present findings with certainty assessments guiding interpretation.

### Subgroup and Sensitivity Analyses

Planned subgroups examine caregiver characteristics (gender, age, relationship), care contexts (setting, disease, region), and study features (design, quality, sample size). Sensitivity analyses exclude high-bias studies, outliers, and test different analytical approaches. Publication bias assessment uses funnel plots and Egger’s test where ≥10 studies enable meta-analysis.

## Discussion

### Anticipated Contributions

This protocol outlines a rigorous approach to synthesizing evidence on caregiver well-being determinants in end-of-life care. The comprehensive scope addresses limitations of previous reviews through inclusive eligibility criteria, systematic quality assessment, and integrated synthesis across methodologies. By employing ecological frameworks, the review will elucidate complex interactions among multilevel determinants. The SPIDER framework ensures appropriate capture of complex caregiving phenomena, while GRADE/CERQual assessments provide transparent evidence evaluation.

Methodological strengths include comprehensive database searching from inception to capture historical development of the field, inclusion of diverse study designs recognizing different contributions to understanding determinants, and use of Covidence ensuring transparent, reproducible processes. The protocol’s emphasis on both positive and negative aspects of well-being addresses previous literature biases toward deficit-focused outcomes. Integration of quantitative and qualitative evidence enables nuanced understanding of both determinant-outcome associations and underlying mechanisms.

### Challenges and Limitations

Several challenges are anticipated. Heterogeneity in well-being conceptualization and measurement may complicate synthesis, addressed through subgroup analyses and careful attention to construct definitions. The complexity of determinant interrelationships requires sophisticated analytical approaches and clear presentation. Cultural variations in caregiving experiences necessitate careful consideration of context in interpretation. Publication bias toward significant findings may overestimate associations, assessed through statistical tests and consideration of null findings.

Limitations include English-language restriction potentially excluding important non-English research, though resource constraints necessitate this decision. Cross-sectional designs predominating the literature limit causal inference, acknowledged in interpretation. The exclusion of grey literature may miss relevant findings, partially addressed through reference searching. The broad scope, while comprehensive, may challenge synthesis depth for specific determinants.

### Implications

Research implications include identifying methodological gaps requiring longitudinal designs examining trajectories, intervention studies targeting modifiable determinants, and research in underrepresented populations. The synthesis will highlight measurement inconsistencies suggesting need for standardized well-being assessment in end-of-life caregiving research.

Clinical implications encompass evidence-based screening tools identifying at-risk caregivers, understanding of intervention targets, and guidance for tailoring support. Healthcare providers will gain frameworks for comprehensive caregiver assessment addressing multiple well-being dimensions.

Policy implications include evidence supporting caregiver assessment requirements, resource allocation for support services, and integration of caregiver needs into quality indicators. The multilevel determinant analysis will inform policies from individual support programs to systemic healthcare reforms.

## Data Availability

As this is a protocol for a systematic review, no datasets were generated or analyzed during the current study. All data included in the future review will be derived from previously published studies and will be cited accordingly. Any extracted data used for analysis will be made available upon reasonable request.

